# Comorbidities might be a risk factor for the incidence of COVID-19: Evidence from a web-based survey of 780,961 participants

**DOI:** 10.1101/2020.06.22.20137422

**Authors:** Mohammad Rahanur Alam, Md. Ruhul Kabir, Sompa Reza

## Abstract

**Background:** The global pandemic of COVID-19 is posing the biggest threat to humanity through its ubiquitous effect of unfathomable magnitude. It has been responsible for over four hundred thousand death worldwide to date. There has been evidence that various comorbidities have a higher risk associated with case fatality. Although COVID-19 is a viral disease, there might be an association between different comorbidities and the occurrence of the disease.

**Method:** Sociodemographic and medical history data on different comorbidities such as asthma, diabetes, liver disease, lung disease, heart disease, kidney disease, hypertension, and obesity were collected by a web-based self-reported survey between 25th March 2020 to 4th June 2020 by the Nexoid United Kingdom. Univariate and multivariate logistic regression analyses were done using these risk factors as independent variables.

**Result:** A total of 780,961 participants from 183 different countries and territories participated in this study. Among them, 1516 participants were diagnosed with COVID-19 prior to this study. A significant risk association was observed for age above 60 years, female gender as well as different pre-existing disease conditions such as diabetes, kidney disease, liver disease, and heart diseases. Asthma and diabetes were the major dominant comorbidities among patients, and patients with existing diabetes were 1.464 (AOR: 1.464; 95% CI: 1.228-1.744), more likely to develop the disease than others who did not diagnose as diseased.

**Conclusion:** Older adults, female as well as people with comorbidities such as diabetes mellitus, heart disease, kidney disease, and liver disease, are the most vulnerable population for COVID-19. However, further studies should be carried out to explain the pathway of these risk associations.

## Introduction

After the emergence of the outbreak of novel coronavirus in December 2019 in Wuhan, China [1], the WHO had announced this outbreak as a Public Health Emergency of International Concern On 30^th^ January 2020 [2]. As of 8^th^ June 2020, there have been almost seven million confirmed cases and Four hundred thousand deaths due to COVID-19 worldwide according to the recent statistics of the World Health Organization [3]. Several scientists have hypothesized about zoonotic transmission, although this is yet to be proven [4].

The Coronavirus infection disease is a highly contagious and transmittable disease caused by a betacoronavirus: severe acute respiratory syndrome coronavirus 2 (SARS-CoV-2) [5]. Coronavirus is a group of positive single□stranded large RNA (+ssRNA) (∼30□kb) viruses [6], containing at least six open reading frames (ORF) and at least four main structural proteins: spike (S), membrane (M), envelope (E), and nucleocapsid (N) proteins [6, 7]. It can infect not only human hosts but also different animals [8]. This virus belongs to the same family, composed of the severe acute respiratory syndrome coronavirus (SARS-CoV), H5N1 influenza A, H1N1 2009, and Middle East respiratory syndrome coronavirus (MERS-CoV) cause acute lung injury (ALI) and acute respiratory distress syndrome [9]. The transmissibility of SARS-CoV2 is higher compared to that of SARS-CoV [10]. Furthermore, the mortality for COVID-19 is higher, but case mortality is higher for MERS-CoV and SARS-CoV [11].

The symptoms of COVID-19 include Fever, Lower respiratory tract infection, acute respiratory infection, Cough, Shortness of breath, etc., [1, 12]. However, there can be COVID-19 case without any signs and symptoms, which makes the control of this pandemics challenging [13]. Geriatric population and people with severe comorbidities such as Asthma, Chronic kidney disease (CKD), chronic lung disease, Diabetes mellitus, Hemoglobin disorders, Immunity deficiency, Liver disease, Serious heart conditions, Severe obesity might be at higher risk of poor clinical outcome [14, 15], increased hospital stays [16] and higher case fatality rate [12]. Along with increasing severity and mortality, these comorbidities might be associated with the incidence of COVID-19, since the immune system and systemic metabolism of the body are already compromised owing to these pre-existing conditions. Our study aims to determine the association between the COVID-19 infection and pre-existing comorbidities using data collected from 780,961 participants from all over the world through a web-based self-reported survey [17].

## Method

A secondary dataset from the Nexoid United Kingdom was used for this study. The data was collected between 25th March 2020 to 4th June 2020 by a web-based self-reported survey. This dataset is under the “Attribution 4.0 International (CC BY 4.0)” license. In order to preserve the confidentiality of the participants, several measures were ensured, such as e-mail address, I.P. address, date of birth was kept private as well as timestamp and location data were randomly adjusted. The survey questionnaire was comprised of 30 questions regarding sociodemographic, behavioral, morbidity, clinical, and diagnostic information related to COVID-19 [17]. A total of 780,961 people filled the online survey, and among them, 1516 people were diagnosed for COVID-19. A survey map was generated using the I.P. location data of the participants using the QGIS application (version 3.4.11) (Figure 1).

**Figure 1:**
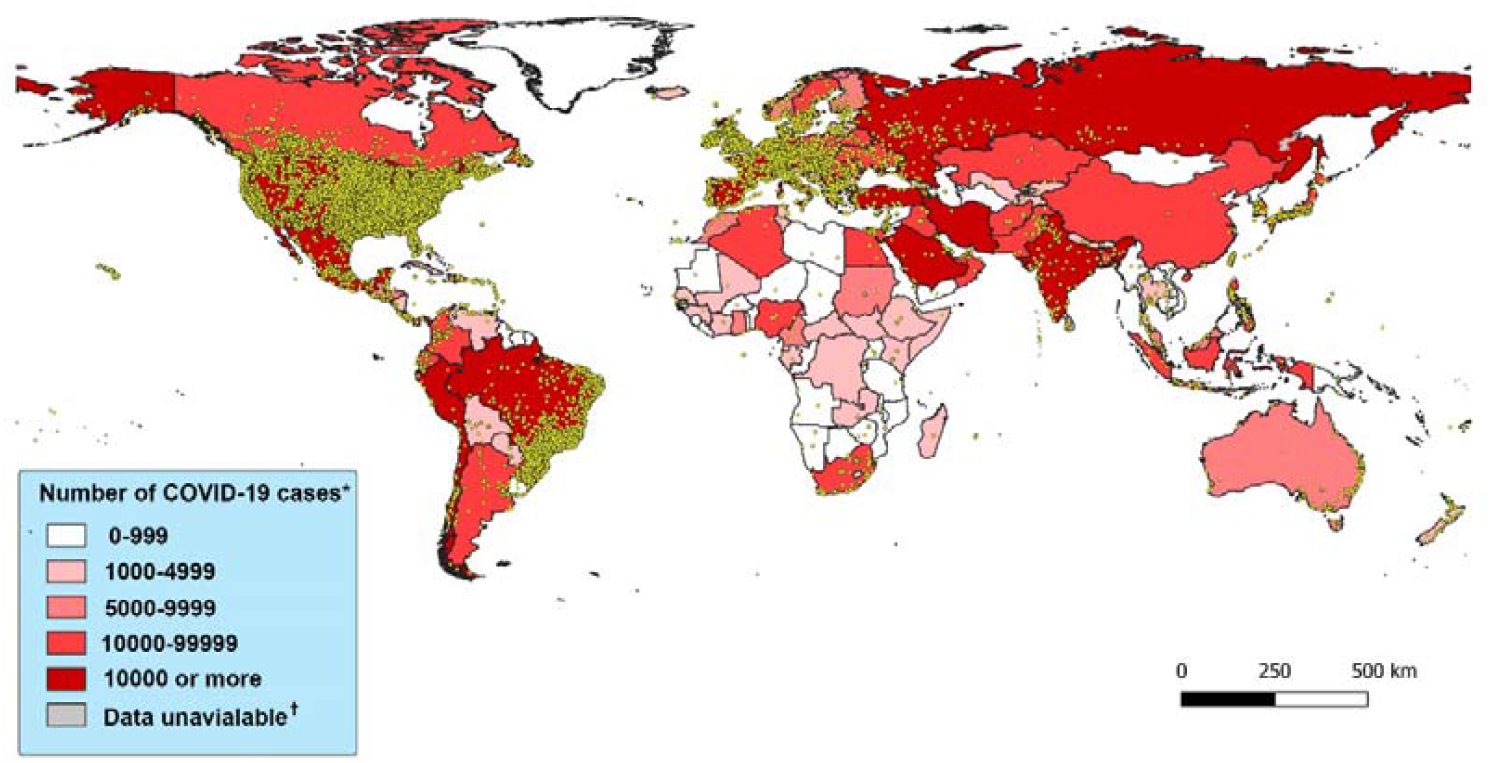
World map according to the intensity of COVID-19 cases and sample location. Note: Yellow dot represents sample locations. * Data were retrieved from worldometers website on 7^th^ June 2020 GMT 18.00 hour [18] † Data was not available for North Korea

Age, Gender, BMI, COVID, and other disease conditions such as Asthma, Liver diseases, Lung diseases, Heart diseases, Diabetes mellitus, Hypertension data were extracted from the database. Obesity was determined from the BMI: People having a BMI of 30 or higher were considered obese.

Data analysis was carried out using IBM SPSS statistics (Version 23). Each variable was tested for descriptive statistics. The Categorical variables are presented as frequency and percentages, where continuous variables are presented as mean and standard deviation, and a t-test was done to compare the mean between COVID 19 cases and control population. Univariate logistic regression was carried out using COVID as a dependent variable and others as independent variables separately. The level of significance for all of the statistical tests was set at 0.05. Finally, each of the significant risk factors from univariate logistic regression was fed into a mixed multivariate logistic regression and odds ratio as well as a 95% confidence interval was calculated., A Hosmer and Lemeshow test was carried out In order to analyze the goodness of fit.

## Result

The majority of the study population are young, and under Forty years of age. Most of the participants are from North America, Europe, and South America. The United States, Brazil, Canada, and Belgium are at the top with the highest number of confirmed cases of COVID-19. The above two-third of the study participants are male, and apart from obesity, the number of other comorbidities is higher in the COVID-19 cases (Table 1).

**Table 1:** Sociodemographic and clinical characteristics of the study population

| Characteristics | Total<br>(n=780961) | COVID-19 cases<br>(n=1516) | Control* (n=<br>779445) | P-value<sup>†</sup> |
| --- | --- | --- | --- | --- |
| Age |  |  |  |  |
| Below 40 years | 463426 (59.34) | 1097 (72.36) | 462601 (59.35) | .000 |
| 40-59.9 years | 251349 (32.18) | 482 (31.79) | 250867 (32.12) | .000 |
| 60-79.9 years | 62886 (8.05) | 143 (9.43) | 62743 (8.03) | .000 |
| Above 80 years | 3296 (0.42) | 76 (5.01) | 3220 (0.41) | .000 |
| Region |  |  |  |  |
| Asia | 6328 (0.81) | 35 (2.3) | 6293 (0.8) | .000 |
| Africa | 1384 (0.17) | 4 (0.26) | 1380 (0.18) | .000 |
| Europe | 41484 (5.31) | 205 (13.52) | 41279 (5.3) | .000 |
| North America | 682258 (87.36) | 1117 (73.68) | 681141 (87.39) | .000 |
| South America | 40623 (5.2) | 144 (9.5) | 40479 (5.19) | .000 |
| Oceania | 8853 (1.13) | 10 (0.66) | 8843 (1.13) | .000 |
| Gender |  |  |  |  |
| Male | 526317 (67.39) | 817 (53.89) | 525500 (67.42) | .000 |
| Female | 251730 (32.23) | 691 (45.58) | 251039 (32.2) | .000 |
| BMI | 29.72 ± 7.92 | 30.29 ± 9.3 | 29.72 ± 7.92 | .005<sup>‡</sup> |
| Comorbidity |  |  |  |  |
| Asthma | 123207 (15.78) | 245 (16.16) | 122962 (15.77) | .000 |
| Diabetes<br>mellitus | 46999 (6.01) | 171 (11.27) | 46828 (6.0) | .000 |
| Kidney Disease | 2625 (0.34) | 46 (3.03) | 2529 (0.32) | .000 |
| Liver Disease | 667 (0.09) | 22 (1.45) | 645 (0.08) | .000 |
| Heart Disease | 13131 (1.68) | 92 (6.06) | 13039 (1.67) | .000 |
| Lung Disease | 10424 (1.33) | 44 (2.90) | 10380 (1.33) | .000 |
| Hypertension | 103883 (13.3) | 273 (18.0) | 103610 (13.29) | .000 |
| Obesity | 275129 (35.23) | 514 (33.9) | 274615 (35.23) | .000 |

Note: The percentages may not add up to 100% due to missing data
\*Participants, who are not diagnosed with COVID-19
<sup>†</sup>P-value for Pearson Chi-Square test
<sup>‡</sup>P-value for independent sample t-test

From univariate logistic regression, Female participants and people age 60 or above had a higher risk of COVID-19. Except for Asthma and Obesity, other comorbidities have shown significantly elevated risk associated with the occurrence of COVID-19 (Table 2).

**Table 2:** Univariate analysis of variable associated with COVID-19 incidence

| Variable | Univariate |  |
| --- | --- | --- |
|  | OR (95% CI) | p-value |
| Age* | 1.82 (1.576-2.101) | .000 |
| Gender† | 1.77 (1.6-1.959) | .000 |
| Asthma‡ | 1.029 (.897-1.180) | .681 |
| Diabetes mellitus‡ | 1.989 (1.696-2.333) | .000 |
| Kidney Disease‡ | 9.426 (7.011-12.673) | .000 |
| Liver Disease‡ | 17.780 (11.59-27.277) | .000 |
| Heart Disease‡ | 3.797 (3.073-4.692) | .000 |
| Lung Disease‡ | 2.215 (1.640-2.991) | .000 |
| Hypertension‡ | 1.433 (1.257-1.633) | .000 |
| Obesity‡ | 0.943 (.848-1.049) | .280 |

\*Age was classified: Below 60 years and above 60 years; Below 60 years were set as a reference for binary logistics regression
†Male gender was set as the reference
‡People who are not diagnosed with the disease were set as the reference

Significant risk factors found in univariate logistic regression were fed into a multivariate logistic regression model. Significantly higher risk associations were observed for Age, Female gender, Diabetes, Kidney disease. Liver disease and Heart disease (Table 3). A Hosmer and Lemeshow test (chi-square: 3.351 and sig .341), confirmed the goodness of fit of this model.

**Table 3:**
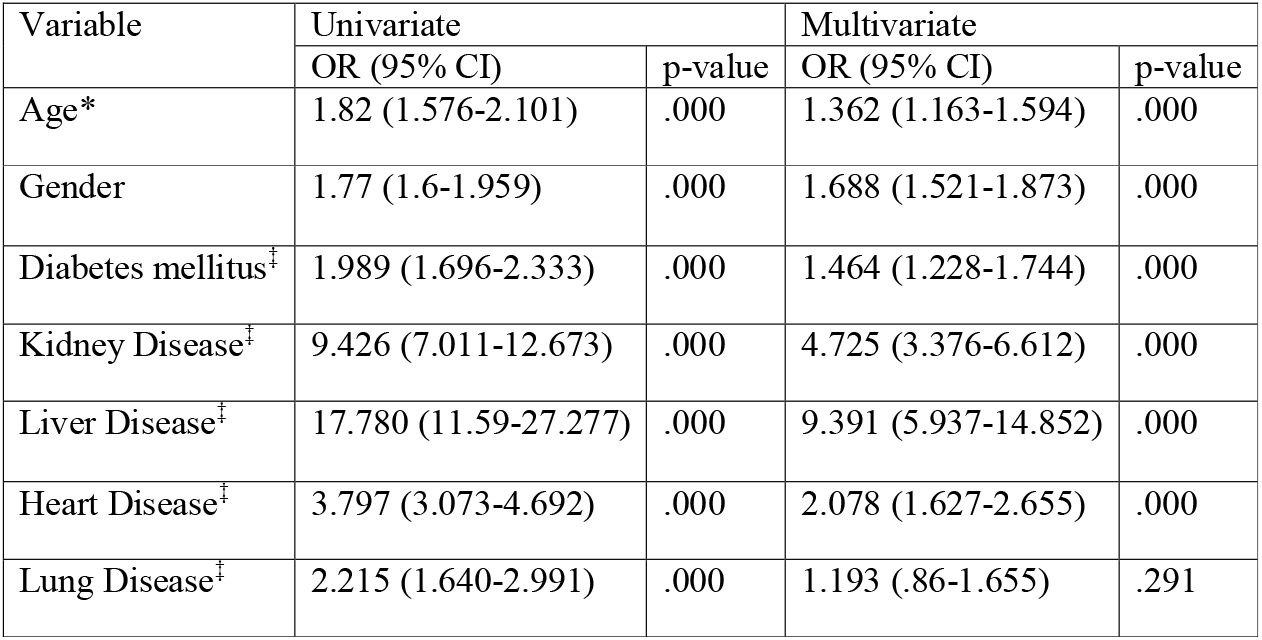
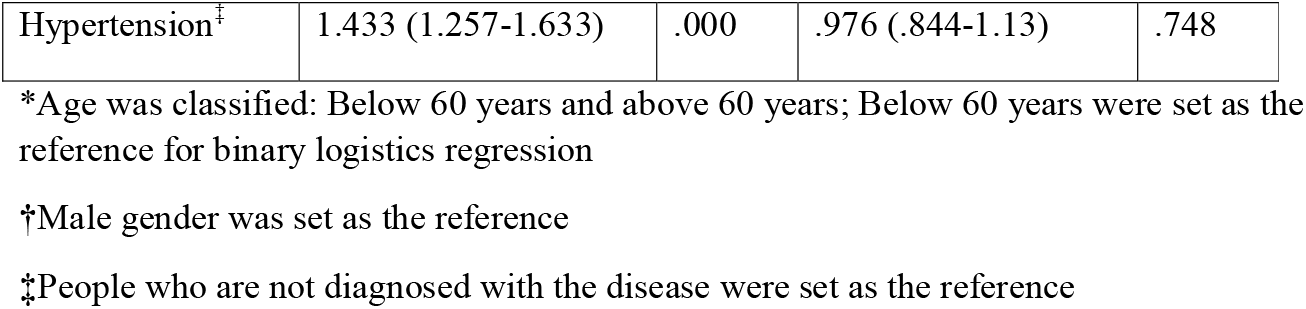
Univariate and multivariate analysis of risk factors associated with the incidence of COVID-19

## Discussion

The present study investigated mostly demographic, morbidity, clinical, and diagnostic information related to COVID-19 from a handful of participants, and among them, 1516 people were diagnosed as COVID positive (+). The study compared COVID positive patients (cases) with controls and found a strong significant association with age, sex, and with critical comorbid situations. Though the majority of the survey participants were male (67.39%), the number of positive cases among females were pretty evenly closed (45.58%) with males. Several studies confirmed that the percentage of men suffering from this virus is higher than women [19], which coincides with our study findings in terms of numbers. The multivariate logistic regression reveals females were 1.688 (OR: 1.688; 95% CI: 1.521-1.873), more likely to get infected by Covid-19 in contrast to male participants. However, this finding does not bode well with one study, which inferred women were less susceptible to this virus than men based on a different innate immunity, steroid hormones, and factors related to sex chromosomes [20]. These contrasting findings must put more debate on this unknown condition in which the world is trying to get an answer. One population-based study conducted in Iceland also found a low incidence in females and children under ten than males [21]

Age was also found to have some effect, and participants over 60 years of age had a marginally high risk (OR: 1.3621.163-1.594) than their younger counterparts though only around 14% of participants were above 60 years. Most of the participants were from the age group under 40 years of age since they must be more used to online surveys and usage. One study conducted in Israel on public data set of 5769 Israeli patients concluded that younger individuals have less chance to experience severe symptoms, which requires intensive care unit hospitalization, and the recovery rate was on average faster than the older aged patients [22]. Another study in China expressed a serious concern, which entails that older age and a high number of comorbidities were associated with higher severity irrespective of gender. However, men’s cases tended to be more critical than women’s [23]

Obesity among participants was seen pretty evenly distributed, and expectedly did not warrant any substantial influence on COVID-19. Hypertension, a common comorbid situation, also assessed to have an insignificant effect on COVID cases after controlling the influence of other predictors. These findings also contrast with one study conducted in Mainland Chain, which found hypertension as the most prevalent (16.9%) comorbidity among 1590 confirmed hospitalized COVID patients [1] and another study where hypertension was even more common clinical manifestation (30%) [1, 24].

To put views on the effect of other comorbidities on COVID-19 in this present study reveals that participants with any of the existing comorbidities (liver, kidney, heart, lung disease, and diabetes) were observed to have more risks of this disease than participants who did not have the conditions. Though the number of COVID cases among kidney disease patients was low, which matches one article published in Nature Reviews Nephrology, stressed that its involvement might result in multiple organ dysfunction and severe outcomes [25]. An understanding of the pathophysiology and mechanisms of kidney damage is still under scrutiny, and effective treatment is still not established [26]. Hence, it should be taken very seriously. Our study reveals that participants with kidney problems had 9.391 times (OR: 9.391; 95% CI: 5.937-14.852) more chance of being infected with the virus than others, and this trend of increased chances were observed with other comorbid conditions though in a lesser degree. Diabetes with another common clinical comorbidity found out in our study coincides with the results of other studies [24]. From the result, it appears that having physiological problems of different kinds had a serious aggravating effect as it increases the likelihood of getting infected by a novel coronavirus. Moreover, having comorbidities also correlated with poorer clinical outcomes in COVID patients [1].

There are some limitations to this study. First of all, this is an online-based self-reported study. Therefore, the accuracy and validity of the data are questionable. Secondly, most of the study participants are from North America, Europe, and South America, but there is a current galloping trend of infection and fatality in Asia. These findings may not apply to the population of these parts of the world. The online survey requires more knowledge, technological skills, and logistics. The people from low and middle-income countries, as well as the older adults, lack the logistics and technological skills for participating in these studies. Finally, kidney disease, lung disease, heart disease, liver disease are composed of an array of diseases. No specific disease was mentioned in this study rather than just groups of diseases. If the particular type of disease was mention that would have provided a more specific and more precise picture.

Though more specific clinical studies are required to understand this unprecedented mystery of a disease and how it affects other deadly comorbidities or whether other comorbidities had any say on this association still requires studies of different kinds to come close to any conclusive remarks. Our study attempted to shed some light on how comorbidities can come in and increases the likelihood of getting infected, and the pathways of infection require clarification, which we believe will be sorted out with further studies.

## Data Availability

The datasets generated during this study are available from the corresponding author on a reasonable request.

## Notes

### Competing Interest Statement

The authors have declared no competing interest.

### Funding Statement

The authors received no specific funding for this study.

### Author Declarations

Ethics approval was not required for this study.

